# Virtual handover of patients in the pediatric intensive care unit during COVID-19 crisis

**DOI:** 10.1101/2021.02.24.21252145

**Authors:** Mohamad-Hani Temsah, Noura Abouammoh, Ahmad Ashry, Ayman Al-Eyadhy, Ali Alhaboob, Fahad Alsohime, Mohammed Almazyad, Majed Alabdulhafid, Reem Temsah, Fadi AlJamaan, Amr Jamal, Rabih Halwani, Khalid Alhasan, Jaffar A. Al-Tawfiq, Mazin Barry

**Affiliations:** College of Medicine, King Saud University, Riyadh, Saudi Arabia; Pediatric Intensive Care Unit, Department of Pediatrics, King Saud University Medical City, King Saud University, Riyadh, Saudi Arabia; Department of Family and Community Medicine, King Saud University, Riyadh, Saudi Arabia; College of Pharmacy, Alfaisal University, Riyadh, Saudi Arabia Division of Pediatric; Critical Care Department, King Saud University Medical City, Riyadh, Saudi Arabia; Evidence-Based Health Care & Knowledge Translation Research Chair, King Saud University, Riyadh, Saudi Arabia; Sharjah Institute of Medical Research, University of Sharjah, Sharjah, United Arab Emirates; Department of Clinical Sciences, College of Medicine, University of Sharjah, Sharjah, United Arab Emirates; Specialty Internal Medicine and Quality Department, Johns Hopkins Aramco Healthcare, Dhahran, Saudi Arabia; Infectious Disease Division, Department of Medicine, Indiana University School of Medicine, Indianapolis, IN, USA; Infectious Disease Division, Department of Medicine, Johns Hopkins University School of Medicine, Baltimore, MD, USA; Division of Infectious Diseases, Department of Internal Medicine, College of Medicine, King Saud University and King Saud University Medical City, Riyadh, Saudi Arabia

**Author notes:** Correspondence to: Dr. Amr Jamal;, PO Box 2925 (Internal Code 34), Riyadh, 11461, Saudi Arabia. These authors contributed equally to this research (Mohamad-Hani Temsah, Noura Abouammoh). Consent for publication: All authors gave their consent for publication. Availability of data and materials: All the data for this study will be made available upon reasonable request. Funding: The authors are grateful to the Deanship of Scientific Research, King Saud University, for funding through the Vice Deanship of Scientific Research Chairs. Conflict of interest: The authors declare no conflicts of interest. Ethics approval and consent to participate: The study was approved by the institutional review board of King Saud University (approval # 20/0553/IRB). Author contributions: MHT, NA, JT, FAJ, AAE, MB, and RH conceptualized the study, analyzed the data, and wrote the manuscript. FAS, AAH, KAH, AA, MAM, MAH, and RT contributed to the study design; collected, analyzed, and interpreted data; and edited the manuscript. NA contribution to the study design and interpretation and edited the manuscript. AJ interpreted the data and finalized the manuscript. All authors reviewed and approved the final version of the manuscript. Author emails and ORCID IDs: Mohamad-Hani Temsah (MHT): PO BOX 2925, Riyadh 11461, Saudi Arabia, Noura Abouammoh (NA): PO BOX 2925, Riyadh 11461, Saudi Arabia, Ahmed Ashri (AA), Ayman Al-Eyadhy (AAE): PO BOX 2925, Riyadh 11461, Saudi Arabia, Ali Alhaboob (AAH): PO BOX 2925, Riyadh 11461, Saudi Arabia, Fahad Alsohime (FAS): PO BOX 2925, Riyadh 11461, Saudi Arabia, Mohammed Almazyad (MAM), Majed Alabdulhafid (MAH), Reem Temsah (RT): PO Box 14135, Riyadh 11424, Saudi Arabia, Fadi Aljamaan (FAJ): PO BOX 2925, Riyadh 11461, Saudi Arabia, Amr Jamal (AJ): PO BOX 2925, Riyadh 11461, Saudi Arabia, Rabih Halwani (RH): PO Box 27272, Sharjah, United Arab Emirates, Khalid Alhasan (KAH): PO BOX 2925, Riyadh 11461, Saudi Arabia, Jaffar A. Al-Tawfiq (JT): PO Box 11705, Dhahran 31311, Saudi Arabia, Mazin Barry (MB): PO BOX 2925, Riyadh 11461, Saudi Arabia.

**Keywords:** COVID-19, PICU, physicians, physical distancing, Zoom for remote handover, tele-ICU

## Abstract

**Objectives:** A key measure to mitigate coronavirus disease 2019 (COVID-19) has been social distancing. Incorporating video-conferencing applications in the patient handover process between healthcare workers can enhance social distancing while maintaining handover elements. This study describes pediatric intensive care unit (PICU) physicians’ experience of using an online video-conferencing application for handover during the COVID-19 pandemic. Design: qualitative content analysis

**Setting:** PICU at a university hospital in Riyadh, Saudi Arabia Subjects: PICU Physicians

**Interventions:** Due to the pandemic, the hospital’s PICU used Zoom^®^ as a remote conferencing application, instead of a face-to-face handover. Following institutional review board approval, data were collected over two weeks (July 1, 2020 to July 14, 2020). Measurements: Demographic data and narrative descriptions of the perceived efficacy of remote handover were collected using open-ended questions through a created online link. The analysis process included open coding, creating categories, and abstraction.

**Main Results:** All 37 PICU physicians who participated in the handover completed the survey. The participants comprised six attendings, nine specialists, and 22 residents. They had variable previous teleconferencing experiences. Most physicians (78.4%) were comfortable conducting a remote endorsement. Most found that Situation–Background– Assessment–Recommendation handover elements were properly achieved through this remote handover process. The perceived advantages of online handover included fewer interruptions, time efficiency, and facilitation of social distancing. The perceived disadvantages were the paucity of nonverbal communication and teaching during virtual meetings.

**Conclusions:** Video-conferencing applications used for online handovers could supplement traditional face-to-face intensive care unit patient endorsement during outbreaks of infectious diseases. The use of video streaming and more emphasis on teaching should be encouraged to optimize the users’ experience.

## Introduction

Since the coronavirus disease 2019 (COVID-19) pandemic was declared in March 2020 (1), healthcare systems worldwide became under unprecedented burden of optimizing care delivery to massive numbers of patients with COVID-19 while protecting healthcare workers (HCWs) from contracting the disease. Furthermore, social distancing measures that were an integral part of the pandemic control needed to be incorporated into the medical care system (2). The rapid spread of the disease, the increasing number of cases and associated mortality, and the lack of therapeutic and vaccine options prompted many governments and health authorities to implement strict measures to combat the pandemic. These include community lockdown, travel and movement restrictions, and cancelation of events and non-essential gatherings (3). Similar to severe acute respiratory syndrome coronavirus (SARS-CoV) and the Middle East respiratory syndrome coronavirus (MERS-CoV), disrupting the chain of transmission is considered key to stopping the spread of severe acute respiratory syndrome coronavirus 2 (SARS-CoV-2). During the evolving pandemic, various strategies should be implemented in the healthcare system and at the local hospital setting (4-7). Healthcare settings can be an important source of viral transmission, and hence, special attention should be given to minimizing the transmission of infections to patients and among HCWs, especially during such a pandemic (8-15).

A key measure to mitigate the spread of SARS-COV-2 has been social distancing to reduce the probability of contact between infected and noninfected individuals (16). The safety of hospitalists and other frontline HCWs is paramount for preventing nosocomial transmission, as reported in several countries (17-19). Much effort to date has focused on maintaining the supply of personal protective equipment (PPE). However, another essential strategy for preventing nosocomial transmission is to implement “physical distancing” and avoid close contact with others. While this approach has received considerable pressure regarding implementation in communities, implementing social or physical distancing measures between HCWs is challenging due to the nature of their job. Still, physical distancing is a critical way of preventing nosocomial transmission and ensuring workforce welfare (20). The Centers for Disease Control and Prevention defined “prolonged” exposure to patients with COVID-19 as a cumulative time of 15 or more minutes in 24 h and close contact as being within 6 ft of a person with confirmed COVID-19 or having unprotected direct contact with infectious secretions or excretions of the person with confirmed COVID-19 (20).

One fundamental hardship of physical distancing is conducting routine clinical care, including rounds, sign-out, or multidisciplinary rounds. Virtual rounds enable clinicians, including residents and attendings, to work together and plan daily care without crowding into patient rooms. This is the most important cultural hurdle that one may face, given the myriad clinical interactions occurring within teams in the hospital; thus, such distancing can be challenging (21).

Effective patient handovers are critical for patient care and safety. This is even more crucial with the restriction of junior doctors’ working hours, resulting in more patient handovers and consequently a greater potential for communication disintegration. The diversity of handover practices with their variable quality and structure can translate to medical errors, treatment delays, and additional tests, resulting to longer hospitalizations and low provider and patient satisfaction (22).

Verbal handovers are often incomplete, with the omission of pertinent information, coupled with poor retention of information by the incoming care provider (23). Electronic handover tools have been reported to help overcome the deficits of the variable and unstructured forms of clinical handover (24).

As multidisciplinary rounds typically occur either at the bedside or in a conference room, promoting to perform them virtually whenever possible during an infectious disease outbreak through either conference calls or video chats is vital (21).

Before the COVID-19 pandemic, the handover process in our pediatric intensive care unit (PICU) between physicians involved all previous on-call physicians and all PICU on-service physicians, with an average of 10–12 doctors sitting together in the PICU nurses’ station to endorse all PICU patients in the early morning. With the COVID-19 crisis and social distancing implementation, a remote handover process was introduced in our hospital on May 14, 2020, where all physicians are in various physical locations in the hospital (Appendix 1).

This study evaluates the feasibility and describes the experience of complete video-conferencing for handover between PICU physicians in a tertiary care academic hospital.

## Methods

### Study design

This study is a qualitative deductive thematic content analysis of the narrative responses from HCWs in the PICU.

### Setting

The physician staffing of the PICU at King Saud University Medical City (KSUMC) consists of six consultants, eight registrars, 4–6 rotating residents from the pediatric department per month, and two PICU fellows. All these physicians, along with nurses, one pharmacist, one clinical dietician, and respiratory therapists, work to serve 15 ventilated PICU beds.

### Sampling and recruitment

All PICU physicians were invited via email to participate in this study. The email was sent on July 1, 2020, with one follow-up reminder after 1 week.

### Data collection

Data were collected online through the SurveyMonkey platform. Open-ended questions were used, and probing was encouraged using questions such as “Why?,” “Can you give an example?,” and “Can you provide details to your answer?.”

The survey started with questions on demographic information (e.g., position, specialty, gender, and age). Then, general questions on identifying obstacles and facilitators of the online handover of care were introduced using probing. Additionally, satisfaction with the Situation–Background–Assessment–Recommendation (SBAR) communication framework was assessed by encouraging the respondents to elaborate and give examples that help understand the identified level of satisfaction of each element.

### Data analysis

The first step in the analysis involved reading and familiarization of the participants’ range of responses. Categories were established, and two authors (NA and MT) developed codes independently. NA, an expert in qualitative methodology working in family and community medicine, introduced an etic perspective of the topic, while MT, a PICU consultant, introduced an emic perspective.

The developed codes were similar and were discussed before a consensus on the coding frame was established. All themes were a *priori* themes; however, the range of responses under each subtheme was derived from the data.

Qualitative data management was conducted using NVivo 10.

After obtaining institutional review board approval, we invited PICU physicians of KSUMC, who have been performing remote handovers using Zoom^®^ since May 15, 2020, to describe their experience through a qualitative, prestructured survey.

Data entry was performed electronically. Content analysis was used to analyze the participants’ responses. The results were used as a part of the quality improvement project and shared with the Pediatric Department Quality Committee.

## Results

Thirty-seven physicians responded to the open-ended questions on using Zoom^®^ for the handover of care between physicians in the PICU (Table 1). The participants comprised six consultants, nine specialists, and 22 residents. Most (86.5%) had previous experience with the implemented Zoom platform (Table 2), and approximately two-thirds also had previous teleconferencing experience in webinars or online learning activities. Furthermore, 78.4% of the participants (n = 29) reported that they were comfortable conducting such handover through Zoom or other similar applications (Figure 1).

**Table 1.** The participants’ demographics.

| Respondent | Position | Specialty | Gender | Age range | Number of previous online endorsements |
| --- | --- | --- | --- | --- | --- |
| 1 | Resident | Others | Female | 31-35 | 5 |
| 2 | Resident | Pediatrics | Female | 26-30 | 3 |
| 3 | Resident | Pediatrics | Male | 26-30 | 5 |
| 4 | Resident | Pediatrics | Female | 26-30 | 1 |
| 5 | Resident | Pediatrics | Female | 41-45 | 9 |
| 6 | Specialist | PICU | Male | 46-50 | 50 |
| 7 | Specialist | PICU | Male | 41-45 | 10 |
| 8 | Specialist | PICU | Male | 31-35 | 10 |
| 9 | Consultant | PICU | Male | 46-50 | 15 |
| 10 | Consultant | PICU | Male | 51-55 | 10 |
| 11 | Consultant | PICU | Male | 36-40 | 14 |
| 12 | Resident | Pediatrics | Female | 26-30 | 6 |
| 13 | Resident | Pediatrics | Male | 26-30 | 2 |
| 14 | Resident | Pediatrics | Male | 26-30 | 4 |
| 15 | Fellow | PICU | Male | 31-35 | 10 |
| 16 | Resident | Pediatrics | Male | 26-30 | 4 |
| 17 | Resident | Pediatrics | Female | 26-30 | 4 |
| 18 | Specialist | PICU | Female | 41-45 | 9 |
| 19 | Consultant | PICU | Male | 36-40 | 1 |
| 20 | Specialist | PICU | Male | 41-45 | 8 |
| 21 | Specialist | PICU | Male | 36-40 | 8 |
| 22 | Resident | Pediatrics | Female | 26-30 | 4 |
| 23 | Fellow | Others | Male | 41-45 | 2 |
| 24 | Consultant | PICU | Male | 31-35 | 7 |
| 25 | Resident | Pediatrics | Male | 31-35 | 4 |
| 26 | Fellow | Others | Male | 31-35 | 1 |
| 27 | Resident | Pediatrics | Female | 41-45 | >10 |
| 28 | Consultant | PICU | Male | 46-50 | 15 |
| 29 | Resident | Pediatrics | Male | 31-35 | 4 |
| 30 | Resident | Pediatrics | Male | 26-30 | 4 |
| 31 | Specialist | PICU | Male | 31-35 | 12 |
| 32 | Resident | Pediatrics | Female | 31-35 | 1 |
| 33 | Resident | Pediatrics | Female | 26-30 | 2 |
| 34 | Resident | Pediatrics | Female | 26-30 | 2 |
| 35 | Resident | Pediatrics | Female | 26-30 | 2 |
| 36 | Resident | Pediatrics | Male | 26-30 | 4 |
| 37 | Resident | Pediatrics | Male | 26-30 | 6 |

**Table 2.** The participants’ previous teleconferencing experience (N = 37).

| Participants' previous teleconferencing experience | n (%) |
| --- | --- |
| Zoom | 32 (86.5%) |
| Online learning | 24 (64.9%) |
| Webinars | 23 (62.2%) |
| Work-related online meetings | 11 (29.7%) |
| FaceTime | 8 (21.6%) |
| Telephone-conferencing | 6 (16.2%) |
| Others * | 2 (5.4%) |
| * Others: Google Meet, Skype, Facebook Messenger |  |

**Figure 1.**
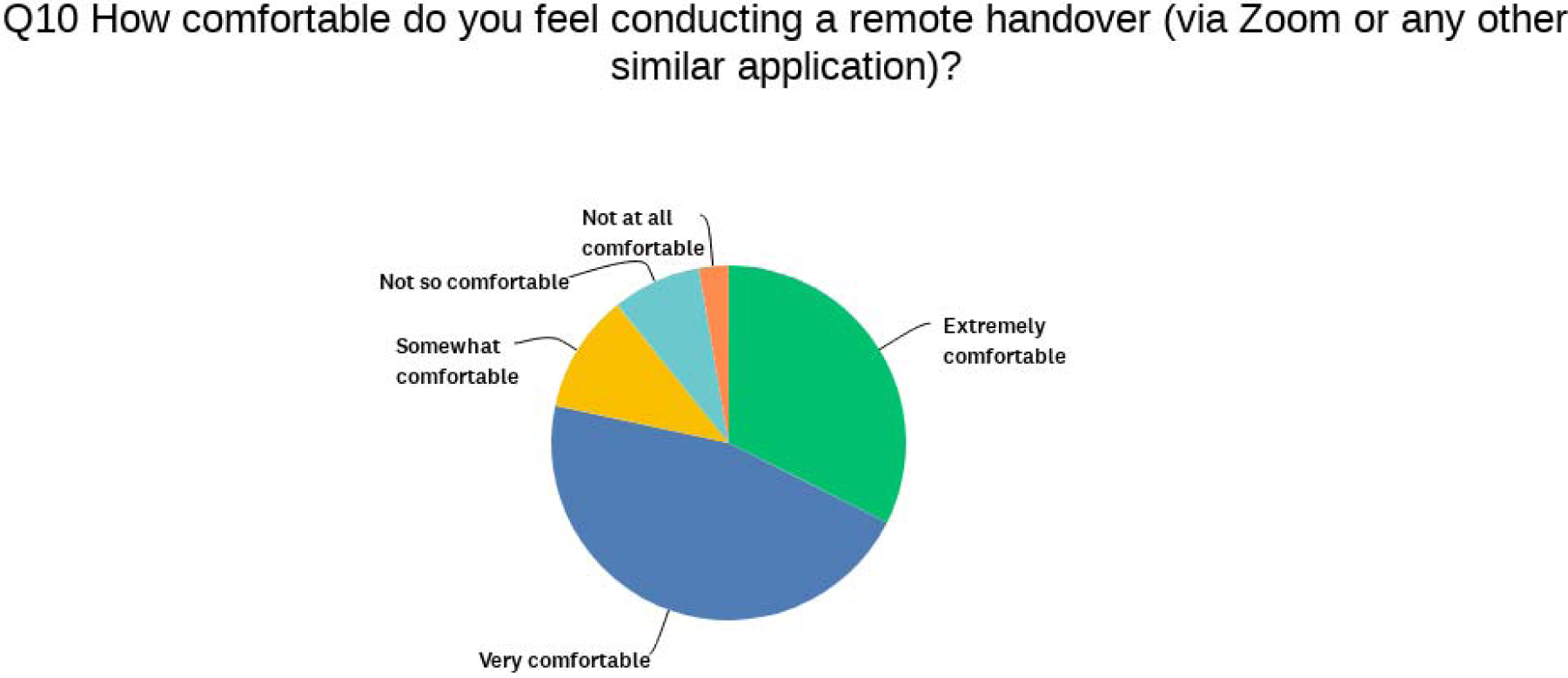
The participants’ comfort level on conducting a remote endorsement (via Zoom or any other similar platforms).

The analysis of the responses revealed two main themes: online and face-to-face endorsement and the influence of online endorsement on quality of care.

### Online and face-to-face endorsement

The participants’ views on the advantages and disadvantages of Zoom endorsement over the classic face-to-face endorsement varied. Most participants found that online endorsement is better than face-to-face endorsement due to time and flexibility, which were also seen by others as factors hindering proper communication among members of the team. For example, Participant 29 who attended four Zoom endorsements noted that *“Zoom endorsement is less time-consuming due to fewer interruptions,”* whereas Participant 25 who attended the same number of online endorsements explained that *“Time is shorter with face-to-face endorsement compared with remote endorsement*.*”* This was explained by Participant 3 who mentioned that *“Sometimes, due to connection errors, it takes more time…face-to-face is usually quicker because there is no disruption of connection and difficulty hearing*.*”*

Regarding the flexibility of use, Participant 22 explained that *“Even if you are late for some reason, you can join the meeting at any place, focusing and writing your own notes*.*”*

Furthermore, physicians who are not on-call may find it easy to participate in Zoom endorsement: *“Sharing information of the patient to all interested (physicians) even not on-call*.*”*

However, some other participants believed that issues including *“less interaction”* and *“less teaching”* are encountered using online methods.

Describing face-to-face endorsement, Participant 5 noted that *“You can see nonverbal communication like facial expression, hand gesture, eye-to-eye contact whereas, with Zoom, you cannot if the cameras are off*.*”* Team members do not use cameras during Zoom endorsement. Participant 4, a resident, noted that *“It wasn’t comfortable as the classical way, in terms of sharing the information without seeing that the other team understood or listened to what you said*.*”* The latter participant had suggested using cameras to increase the reliability of Zoom endorsement. In addition, Participant 4 noted that *“Video calls between teams would be more comfortable*.*”* However, on their practice of sharing images as needed during Zoom endorsement, Participant 7 commented that *“I can share data as X-ray reports, and many subspecialties can attend and share their experience*.*”*

The participants were concerned about teaching, and they commented on that as part of the attributes of working at the PICU in a university hospital, as Participant 18, a PICU specialist, mentioned that *“Teaching of residents is minimal during endorsement*.*”* However, few participants observed no difference in using both manners.

### Influence of online endorsement on quality of care

Responses revealed the participants’ views on the effect of online endorsement on the quality of patient care and endorsements.

### Quality of patient care

Although some participants (n = 12) believed that the quality of work and patient care was not affected by online endorsement compared with face-to-face endorsement, others perceived some positive (n = 15) or negative (n = 9) effects.

The quality of the work environment and patient care was maintained using Zoom endorsement. Participant 20 mentioned that *“Remote endorsement has achieved the main goal of social distancing. Optimum patient care is being achieved with a smaller number of healthcare workers, attending consultant can attend the handover; so earlier decisions could be offered*.*”* Participant 32 added that *“I believe it (Zoom endorsement) will definitely improve patient care. Since I will get the information I need without having to worry about running late and missing some information/updates about the patient*.*”*

In addition to *“less noise,”* Participant 28 believed that not conducting the endorsement at the bedside decreased the interruptions from individuals outside the PICU team during rounds and reported that *“One advantage of remote endorsement is the avoidance of interruptions from family members or other teams which could happen in face-to-face endorsement*.*”*

Considering the COVID-19 pandemic, Participant 9, a consultant, noted the following about the quality of the work environment due to online endorsement: *“Decreasing the stress among HCWs during COVID-19 crisis and decreasing the possible contact with other asymptomatic SARS-COV-2 carriers. So, we have a more sustainable healthcare workforce to take care of more PICU patients*.*”*

Alternatively, some participants believed that *“missed information and discussions”* are commonly observed during online endorsement. However, three participants observed that revising the patient’s status is the senior’s responsibility, regardless of the information shared during the endorsement. Participant 3 noted that *“…I don’t think it does (effect patient care) because the PICU seniors always revise their patients thoroughly*.*”*

Participant 5, a resident, felt that sometimes, the low online attendance might negatively affect the quality of patient care and noted that *“Less number of Zoom attendees will have a smaller number of doctors who will critique or ask questions during the endorsement, less chance for the exchange of views and opinions*.*”*

Furthermore, clinical assessment of the patient can be compromised due to remote endorsement. For example, Participant 15 said that *“Most of the physicians will look all the time to a computer for endorsement and patient follow-up without assessing patients clinically*.*”*

While accepting the adverse effects of online endorsement on the overall work environment, some physicians denied this effect on patient care. Participant 27 noted that *“With the constraints enumerated above, there is always the possibility of miscommunication*.*”* However, when asked if that affects the quality of patient care, Participant 27 responded that *“Patient care is the same with the face-to-face endorsement*.*”*

Connectivity issues have further influenced the quality of patient care. Participant 3, a resident, mentioned that *“Sometimes, there are active patients and the time is not enough; we once had a patient who was a case of pulmonary embolism, she was very active, 85% of the time was spent talking about her condition, and we went over the other patients very quickly because of Zoom limited connectivity issues*.*”*

### Quality of online communication

The descriptive analysis of the SBAR communication tool is shown in Figure 2. Among the 37 participants, 13 provided details based on which they have chosen their SBAR communication tool evaluation.

**Figure 2.**
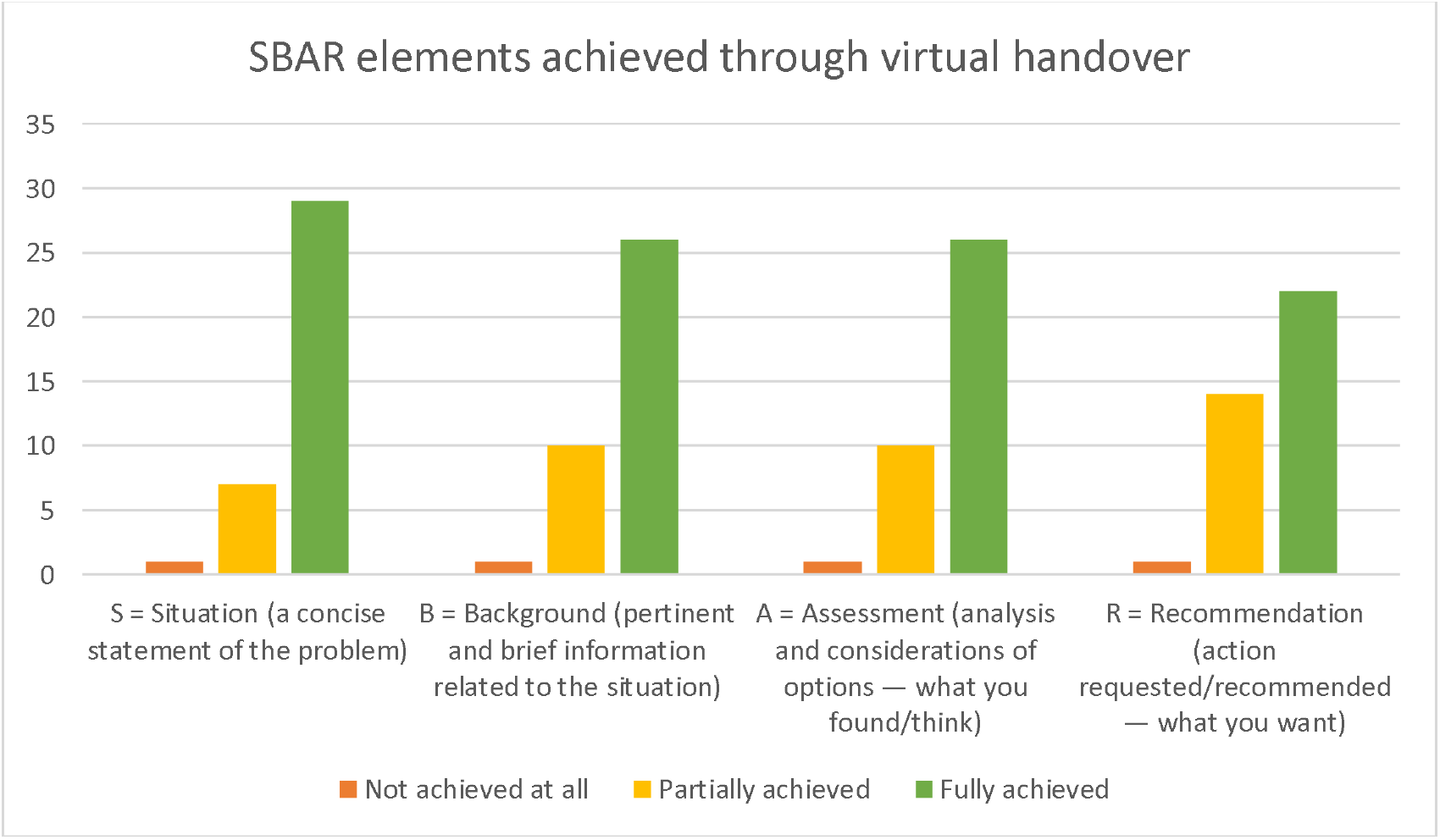
The participants’ responses to achieving quality SBAR elements using online endorsement.

Most participants were satisfied with the quality of communication using online applications, whereas others were less satisfied with the quality of communication for various reasons. The following shows the participants’ responses to each element of the SBAR communication tool.

### Situation

Most participants agreed that the explanation of the situation of each patient endorsed during online endorsement was clear and explicit. Participant 8, a PICU specialist, mentioned that *“We simply can describe the situation to all registrars and residents and also to consultants in their homes*.*”*

Some other participants noted no difference between the classic face-to-face endorsement and the online one, as the latter does not influence the endorsement contents. Participant 28, a PICU consultant, noted that *“It’s (the situation element) fully achieved because it’s the way we used to do it, either face-to-face or remotely*.*”* Alternatively, few participants believed that providing a concise statement of the problem during endorsement is overestimated, as Participant 27, a resident, noted that *“The rotator will usually present the case, and if the PICU on-call feels that information was not enough, he/ she fills the gap*.*”*

### Background

Most participants believed that the patients’ background information is well communicated during online endorsement and understood the importance of this element, especially during the high turnover of patients due to COVID-19 admissions. For example, Participant 4, a resident, explained that *“Because of the new COVID situation, some of the team members would not be aware what happened to the patient last week except through this Zoom meeting…”*

The participants did not observe a difference in the quality of endorsing background information between online and face-to-face endorsements. Participant 32 mentioned that *“Detailed background could be obtained through remote handover”* Participant 20 *“The background information of the patient is the first thing we mention during endorsement, so it is well achieved*.*”*

Nevertheless, few participants thought that a high-quality sharing of background information was partially achieved due to the following: Participant 28 mentioned that *“It’s (background information of the patient) partially achieved because sometimes we lose attention and get some background information lost. While in face-to-face, we are less likely to be distracted*.*”* In addition, Participant 31 noticed *“Some missed information*.*”* Participant 31 believed that sharing background information was partially achieved and had not put the responsibility to fill the missing information on the on-call team.

### Assessment

Like the aforementioned SBAR tool elements, most participants believed that the assessment findings of the patients were well communicated via online endorsement. Participant 3 said that *“(Patient) assessment of the previous team is always mentioned*.*”*

Furthermore, the participants explained some reasons that facilitated the perceived quality of communication of the team assessment of the patients. For example, Participant 9, a consultant, stated that *“Assessment was clear and backed with some radiological images or other documents sharing through (Zoom^®^ platform)*.*”*

However, some participants believed that communicating the assessment element is difficult to achieve through online endorsement as this depends heavily on in-person clinical evaluation. For instance, Participant 20, a specialist, noted that *“Proper assessment needs more than virtual handover, needs personal assessment and clinical examination*.*”*

### Recommendation

Some participants were satisfied with the quality of communication of recommendations during Zoom endorsement. For example, Participant 3 noted that *“Plans and what to follow-up are clear*.*”*

Moreover, another participant added that consultants’ availability to inform the team about and discuss recommendations is essential and easy to maintain during online endorsement. Participant 8, a specialist, mentioned that *“We can take recommendations directly from the consultant remotely*.*”*

Alternatively, Participant 20, a consultant, believed that the elements of the SBAR communication tool are connected. As he believed that a proper assessment is lacking, he noted according to his experience that *“Due to incomplete assessment, an effective plan couldn’t be suggested properly*.*”*

Furthermore, some participants believed that most recommendations in the PICU do not require optimum communication skills, as Participant 32, a resident, mentioned that *“This is partially achieved because we don’t really request anything during endorsement other than mentioning that the patient should be seen by neurology team, for instance*.*”*

Noteworthy, since the initiation of this remote handover process among the PICU physicians from May 15, 2020 to February 15, 2021, contact tracing in our PICU infection control reported only one physician who had a COVID-19-positive polymerase chain reaction result, which was community-acquired. Incidentally, among the other PICU team members who are not using the remote endorsement process, seven nurses (3 of them nosocomial) and two respiratory therapists (one nosocomial) were COVID-19 positive during this period.

## Discussion

As part of the preparation for the COVID-19 pandemic, lessons from similar outbreaks have helped establish known preventive measures; infection prevention and control strategies were developed and revised as the pandemic evolved and transmission and exposure information grew. As part of hospital preparedness for the large influx of infected patients, healthcare facilities were challenged with the limited number of airborne infection isolation rooms and intensive care unit (ICU) beds (25, 26). Hence, additional measures were introduced within hospitals, including universal masking and social distancing. Although social distancing has been a well-established mitigation measure in community settings (16), its effects were relatively unknown within hospital settings. Emerging evidence at the beginning of the pandemic showed that social distancing was a vital component of infection prevention to reduce nosocomial outbreaks. Subsequently, many academic hospitals implemented social distancing during educational and administrative meetings, clinical workrooms, rounds, sign-out, and multidisciplinary rounds. Subsequently, public health authorities issued guidelines for COVID-19 infection prevention in the workplace, which emphasized social distancing (27, 28).

Tele-critical care reduces cost and improves the quality of care using low-cost, off-the-shelf, synchronous, video-teleconferencing devices, along with remote access to electronic medical records, imaging studies, and lab results (29). Video-conferencing technologies, such as FaceTime^®^, Zoom^®^, and Skype^®^, were utilized to assist in family discussions and goals of care settings at the end of life in PICUs during the COVID-19 pandemic (30). Therefore, we theorized that implementing a similar system during the pandemic could maintain the standard of ICU patient care while enabling more social distancing measures.

The handover of patients is critical of any hospital care, especially in settings with complex patients when multiple professions contribute to patient care.

In addition, unambiguous and precise communication is provided by face-to-face communication (31). However, virtual endorsements using applications, such as Zoom^®^, make attending a session easier. Alternatively, low attendance might be of concern when using this technique.

A study has shown unanimous satisfaction of the participating neonatologists, nurses, and the infection control team (32). In one study, physicians felt that their clinical decisions might be negatively impacted by inappropriate health information using online tools (33). The satisfaction of the team involved is of immense help to sustain and improve virtual handover.

One of the participants in this study highlighted the avoidance of family members during the pandemic. However, note that the patient’s or family members’ presence may enrich the handover as they provide valuable input (34). Virtual huddles to enhance staff communication about patients had been used in ICUs during the COVID-19 pandemic. However, it was feared that the speedy adaptation of virtualization might pose the risk of decreasing the quality of clinical care (35, 36). On the contrary, one study has shown that virtual programs may provide additional inpatient capacity during the COVID-19 pandemic (37).

Most participants in this study were comfortable using Zoom^®^ or other similar applications in the handover process, which could be related to their previous experience in using these tools. Lowe and Shen have reported their emergency department’s rapid adaptation of telemedicine network using off-the-shelf products with Apple iPads running Zoom, a familiar system for end-users for physical distancing, reducing high-risk contacts and conserving PPE (38).

However, the use of video cameras throughout the handover process could have intensified the team’s reliability and engagement. This needs further exploration and emphasis, as body language is an integral component in the communication process. Paying attention to the types of nonverbal communication in face-to-face handovers and educating HCWs could improve the quality and reliability of these practices (39).

In this study, the participants find communication and elaboration difficult in online endorsement, which may make achieving a quality online endorsement difficult; however, the so-called electronic ICUs have been established during the COVID-19 crisis to enable clinicians to monitor the clinical status of up to 100 patients spontaneously, provide them rapid access to subspecialty consultation, and allow the continued ability of quarantined staff to continue their work remotely (40).

Even though most participants were post-graduate residents, one possible limitation is their high turnaround, and they may not grasp the whole experience. Implementing virtual handover and telemedicine at other clinical areas within the scope of their rotations may provide additional experience and perspective, which should be further evaluated. Residents’ and trainees’ opinions on Zoom’s use in the clinical practice during the COVID-19 crisis were wildly varied, which is common in narrative analyses (41). A recent cross-sectional survey that evaluated post-graduate residents’ knowledge on infection prevention and control practices did not show any difference in overall knowledge by age, residency year, or rotating department (42).

Some of our training residents thought the whole endorsement process does not affect the patient care in the PICU, thinking that this relates more to the PICU senior staff themselves rather than relating to the trainees. This misconception highlights that training residents need more education on how proper endorsement affects the quality of patient care and safety (43). Having all PICU team members share the handover information about a patient’s current situation, assessments, and care recommendations could prevent near misses and adverse events (43). Unfortunately, such vital information does not always pass flawlessly from the previous to the subsequent healthcare providers. The Agency for Healthcare Research and Quality has reported several gaps in communication between healthcare providers as the leading cause of preventable medical errors in malpractice claims affecting emergency physicians and trainees (44).

Some pieces of evidence exist on the effectiveness of SBAR implementation on patient outcome, but this evidence is limited to specific circumstances, such as communication over the phone (45). As high-quality studies are still lacking, future studies are needed to demonstrate the benefit of SBAR in patient safety and keep raising the awareness of communication errors. SBAR might be an adaptive tool suitable for many healthcare settings when clear and effective interpersonal communication is required (45).

The current COVID-19 pandemic provides various opportunities for using remote communication to develop healing human relationships. What we need in a pandemic is not social distancing, but rather physical distancing with social connectedness (46). The inherent purpose of a multidisciplinary conference (MDC) is to ensure a thorough evaluation of each case, regardless of the spectrum of care, whether pretreatment, treatment, or survivorship (47). This entails ensuring proper diagnosis, staging, treatment planning, clinical trial enrollment, care coordination, management of treatment complications, evaluation of disease response, recurrence monitoring, and assessment of survivorship outcomes. As MDC usage has become more widespread, academic institutions are beginning to evaluate MDC quality measures for guideline adherence and patient outcomes. Given its impact, MDC has become a standard in pediatric critical care.

A virtual MDC makes attendance easier, particularly for off-site healthcare providers. This encourages greater participation for community healthcare providers. The virtual format provides flexibility for on-site healthcare providers as well, promoting attendance. Images are more easily viewed by both neuroradiologists and MDC participants when a virtual format is employed. Furthermore, by attending an MDC at their workstation, healthcare providers have real-time access to patient records, which can be reviewed to assist with clinical decision making.

Family-centered care is threatened during the COVID-19 pandemic (48). The participation of family members in a manner that allows families, patients, and the healthcare team to collaborate is the core of family-centered care. Strategies for delivering family-centered care typically include the open presence of family members at the bedside. Restrictions on family presence should not undermine adherence to the principles of family-centered care. Defining patients’ goals of care is a priority during the pandemic and typically necessitates family engagement. Therefore, rapidly adapting family-centric procedures and tools is essential to circumvent restrictions on physical presence. During the COVID-19 pandemic, family presence must be supported in nonphysical ways to achieve family-centered care (48).

One limitation to our research is the subjective component inherent to this study design. Another issue is the single center experience, so this could be further explored in multicenter trials on utilization of similar remote handover applications in the ICUs.

## Conclusions

Video-conferencing applications used for online handovers could supplement traditional face-to-face ICU patient endorsements during infectious disease outbreaks. The utilization of video streaming and more emphasis on teaching should be encouraged to optimize the users’ experience.

## Data Availability

All the data for this study will be made available upon reasonable request.

## Abbreviations

CDC: Centers for Disease Control and Prevention
COVID-19: coronavirus disease 2019
ICU: Intensive care unit
KSUMC: King Saud University Medical City
MDC: multidisciplinary conference
PCR: polymerase chain reaction
PICU: pediatric intensive care unit
PPE: personal protective equipmentSARS-CoV-2, severe acute respiratory syndrome coronavirus 2
WHO: World Health Organization

### Appendix 1

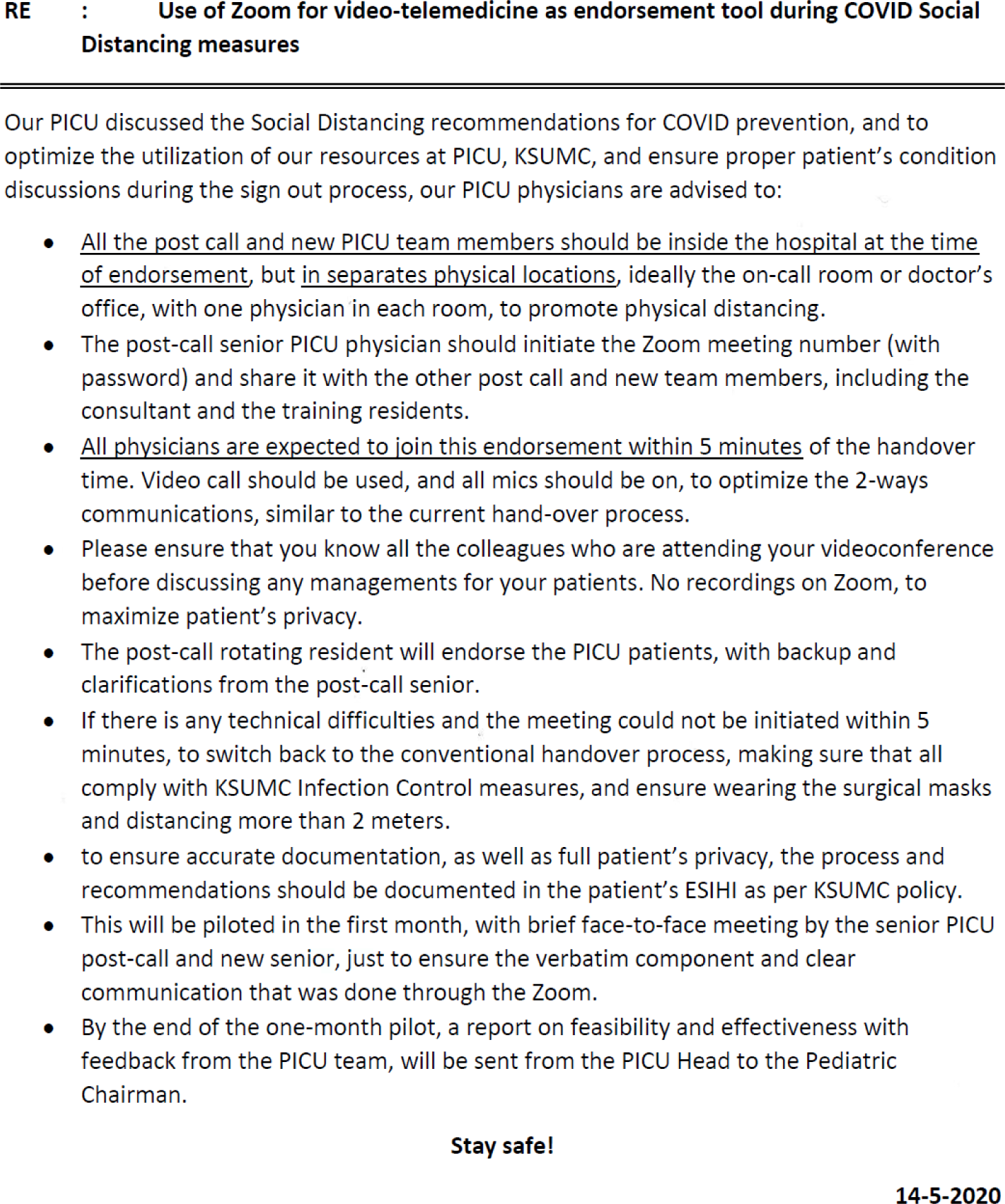

## Notes

### Competing Interest Statement

The authors have declared no competing interest.

### Author Declarations

The study was approved by the institutional review board of King Saud University, Riyadh, Saudi Arabia (approval # 20/0553/IRB).

